# Auto-METRICS: LLM-assisted scientific quality control for radiomics research

**DOI:** 10.1101/2025.04.22.25325873

**Authors:** José Guilherme de Almeida, Nickolas Papanikolaou

## Abstract

The quality and integrity of scientific publications in clinical machine-learning is of paramount importance. Low-quality publications can offer excessively optimistic estimates of model performance, leading to unrealistic expectations when translating models to clinical scenarios. In radiomics this is no less the case: recent studies have highlighted the stark publication bias in radiomics research, and multiple investigations showed how radiomics-based research can be affected by several methodological confounders. To address this systematically, the Radiomics Quality Score (RQS) and the METhodological RadiomICs Score (METRICS) were designed. Both provide a standardised quantity determining the methodological quality of radiomics scientific manuscripts, thus validating their clinical translation. METRICS, out of the two, has been shown to be more reproducible and accurate. In recent years, large language models (LLMs) have been used in many science-related tasks. Here, we ask the following question: how are LLM-based METRICS assessments correlated with those performed by radiologists? Using two recent reproducibility studies, we provide evidence that LLM-based radiomics assessments can be a useful assistant in determining the scientific quality of radiomics-based publications. Particularly, we show that inter-rater agreements between LLMs and human raters are similar to those reported between human raters. Additionally, we also show that the correlation and error between METRICS scores obtained by human raters and LLMs is similar to those obtained between human raters. These results constitute an important proof of concept — LLMs can be used to assist human raters in deriving standardised scores.

**Keypoints:** *Question:* How well do LLM-based METRICS assessments correlate with those performed by radiologists in determining the scientific quality of radiomics-based publications, addressing the need for efficient quality control?

*Findings:* LLM-based radiomics assessments demonstrate inter-rater agreements with human raters similar to those between human raters, suggesting their utility in assisting with standardized scoring.

*Clinical Relevance Statement:* LLMs can assist in the standardized assessment of radiomics research quality, potentially improving the reliability and clinical translatability of radiomics-based tools by ensuring studies meet rigorous methodological standards, ultimately benefiting patient care through more robust clinical applications.

## Introduction

Machine-learning (ML) studies can suffer from methodological biases or errors which inflate their conclusions [1]. Coupled with fast-paced innovation in the world of artificial intelligence (AI), this can lead to excessive confidence in what AI and ML can achieve [2]. In clinical ML this is no different: when analysing methodological pitfalls in the literature, Maleki and others showed that three major ones led to unrealistic performance overestimates in head and neck computed tomography (CT), lung CT, chest radiography and histopathology data [3].

Radiomics — the subfield of clinical ML focusing on extracting patterns from radiology images — has also been shown to be affected by similar biases. Importantly, radiomics literature tends to be biased towards positive results, thus inflating the perception of the efficacy of these methods [4]. Lu and others showed that such conclusions can be attributable to uncontrolled confounders such as the inclusion of features associated with slice thickness or tumour size [5]. Canella and others showed how different data splits could lead to serendipitous performance estimates in microvascular invasion prediction in hepatocellular carcinoma [6]. Demircioğlu showed how the application of different feature selection strategies (typically recommended in radiomics research) influences performance [7]. To further complicate this, CT acquisition parameters can largely make radiomic features impossible to reproduce [8]. In 2016, Gillies and others claimed that, for radiomics, “images are more than pictures, they are data” [9]. Years of poorly designed publications have, however, drawn the attention to a competing (albeit confusing) claim by Pinto dos Santos and others [10]: “are images really data or just patterns in the noise”?

Indeed, the application of radiomics to the clinic requires that methods are better assessed as a segue to make conclusions more realistic [11,12]. To try to make radiomics research more robust and easier to analyse, two distinct radiomic quality metrics were developed in recent years: the Radiomics Quality Score [13] and the METhodological RadiomICs Score (METRICS) [14]. While both are relatively straightforward, a recent comparison of both showed that the latter is more reproducible and accurate [15,16], having motivated a number of studies which apply it to radiomics studies. Two of these studies — Akinci D’Antonoli and others (2025) [17] and Kocak and others (2025) [16] focus on the reproducibility of METRICS. Their excellent reporting provides individual answers for the 30 questions available in METRICS for 6 reading conditions and 3 raters, respectively.

Large language models (LLMs) are recent technological advances characterized by the remarkable capacity of generating human-like text. Their application on several different topics such as molecular property prediction [18] or predicting experimental results [19]. One such domain is that of assessing scientific manuscripts, where LLMs have been successfully used to analyse scientific summaries [20], provide feedback for scientific manuscripts [21], screen systematic reviews [22], and potentially increase review quality by providing feedback [23].

Here, we make use of the recent literature on METRICS reproducibility to answer a simple question: are inter-rater agreements between LLMs similar to that observed between expert raters? To answer it, we use a simple protocol which combines extensively detailed prompting with structured output generation. We analyse the inter-rater agreement between radiologists and groups and a commercially available LLM, showing how differences between human raters are similar to those observed between human raters and the commercially available LLM. For ease of dissemination, we make a small demo available at auto-metrics.netlify.app.

## Methods

### Data collection

We collect the answers for all 30 items and 5 conditions used in METRICS and described in [14] from two different sources: Akinci D’Antonoli and others (2025) [17] and Kocak and others (2025) [16].

We collect the data from Akinci D’Antonoli and others (2025) available as Supplement 1 in the original publication [17]. These data are characterized by 6 distinct conditions: 2 treatment conditions (with and without training on how to use METRICS as a quality assessment tool) and 3 levels of expertise (1, 2 and 3, which take both years practicing and academic experience in producing and evaluating radiomics studies). These conditions were populated by 12 different radiologists, which were equally divided into one of two treatment conditions (received/did not receive training) and into one of three levels of expertise. We systematically retrieved the text for all 34 publications by copy-pasting it from the article webpage, having excluded 3 to which we had no access to the full-text.

For Kocak and others (2025), which analyses specifically articles on radiomics analyses of glioma, we retrieved the individual METRICS scores from three supplementary figures, one provided for each radiologist [16]. Each radiologist annotated 27 articles. We were able to retrieve 22 full-text articles similarly to what was performed for Akinci D’Antonoli and others (2025).

The complete list of articles, annotated for whether or not they were retrieved is presented in Supplementary Table 1.

### Large language model experiment

We use, for all experiments, Gemini Flash 2.0 accessed through its API on March 31st, 2025. A temperature of 0.0 was used to ensure reproducibility and consistency. Below we detail our approach, which we call “Auto-METRICS” (Automated METRICS). The choice of this particular model was purely heuristic — while it is true that different models are likely to yield different results, we focused on a single model as this allows us to showcase whether LLMs have the potential to assist in automatic standardised assessment of scientific production.

### Prompt construction

To construct the prompt we use the prompt detailed in the Supplementary Methods. In short, we provide:

1. Instructions to the prompt, explaining how it should analyse the text of each manuscript;
2. Details on what METRICS is, what it tries to analyse and some of its particularities (i.e. 30 questions and 5 conditions which will define whether some questions can be answered)
3. The evaluation rubric for each question/condition: choose between yes, no and n/a (if a condition allows a question to be skipped) and provide a reason for each evaluation
4. The input format: an entire scientific article
5. The output format: a structured output format containing a summary for the scientific publication and the answers to each of the 30 questions and 5 conditions.

The prompt was further expanded to include the complete guidelines available in the METRICS online tool (https://metricsscore.github.io/metrics/METRICS.html, accessed on March 30th, 2025) [14]. We copied all of the items and conditions as well as the explanations available in each tooltip to further expand and clarify what each item and condition represented in free-text format.

### Constrained generation

We used constrained generation (i.e. structured output) generation as made available from the Google Gemini API [24]. This required the specification of 30 items and whether or not their answers are dependent on any given condition.

### Implementation

We make all of the code used for this manuscript in github.com/josegcpa/auto-metrics. Shortly, we used Python version 3.13 and Google Gen AI Python SDK (google-genai) version 1.8.0 to perform LLM queries to Gemini Flash 2.0 and Pydantic version 2.11.1 to specify the structured output format. We make a small web app available in auto-metrics.netlify.app that allows users to freely use Auto-METRICS with their own Google AI Studio API keys.

### Prompt enhancements

We first tested Auto-METRICS on the scientific articles analysed in Akinci D’Antonoli and others (2025). Upon noticing that the Gemini 2.0 Flash output was systematically incorrect for specific items, we further improved the explanations for each to better improve this. We then applied the improved methodology to Kocak and others (2025), using this to validate our prompt. The protocol to do this consisted in analysing manuscripts where the average agreement between the LLM and the raters was lowest (the agreement was 1 when the LLM and the human rater group agreed, 0 if the item was not considered by either, −1 when they disagreed).

We term the initial LLM prompt outputs as “LLM” and the enhancements as “LLM + changes”.

## Statistical analysis

To compare different raters (human groups, human or LLM-based) we make use of Cohen’s Kappa and use it as a measure of inter-rater agreement. We make comparisons stratified at three different levels: i) considering all items and conditions, ii) considering the average Cohen’s Kappa across items and iii) considering the average Cohen’s Kappa across conditions. These estimates are compared with one another using their confidence intervals, considering that statistically significant differences exist whenever two confidence intervals calculated at 95% are non-overlapping. To calculate 95% confidence intervals for the average Cohen’s Kappa calculated across items/conditions, we consider the standard error of the estimate and use it to calculate the upper and lower bounds of the estimate (approximately this is 1.96 * SE, where SE is the standard error).

A key aspect of METRICS is that it allows for an overall score of quality which consists of a weighted average of all items. We calculate this for all estimates and perform an ANOVA for the annotator label (8 groups for Akinci D’Antonoli and others (2025) and 5 groups for Kocak and others (2025)) followed by post-hoc Tukey’s Honest Significant Difference tests if the ANOVA was statistically significant.

Both analyses are performed independently for both reproducibility studies.

## Code and data availability

We make all code (for generation using LLM and analysis) and data (article texts, inter-rater agreements as extracted from the paper, prompts, and LLM generations) available in https://github.com/josegcpa/auto-metrics. To assist researchers in using this method, we provide an easily accessible website at https://auto-metrics.netlify.app/.

## Results

### Similarities between large language models and human raters

As visible in Figure 1 and highlighted in the original publication [17], there is limited interrater agreement between human rater groups. Importantly, there is one considerable exception — the value for Cohen’s Kappa measured between expert raters with and without training is considerably higher than what is observed for other groups (⊠=0.79 for the entire METRICS questionnaire, 95% CI=[0.75, 0.83]; Figure 1**A**). When analysing the average interrater agreement for items and conditions, this is also the case: the experienced rater groups with and without training rated scientific publications similarly (Figure 1**B,C**). In general and excluding comparisons between experienced rater groups, the observed scores align with those reported in the original publication, i.e. moderate agreement. Within individuals with training, we observe higher levels of agreement, highlighting how training can improve the reproducibility of METRICS.

**Figure 1.**
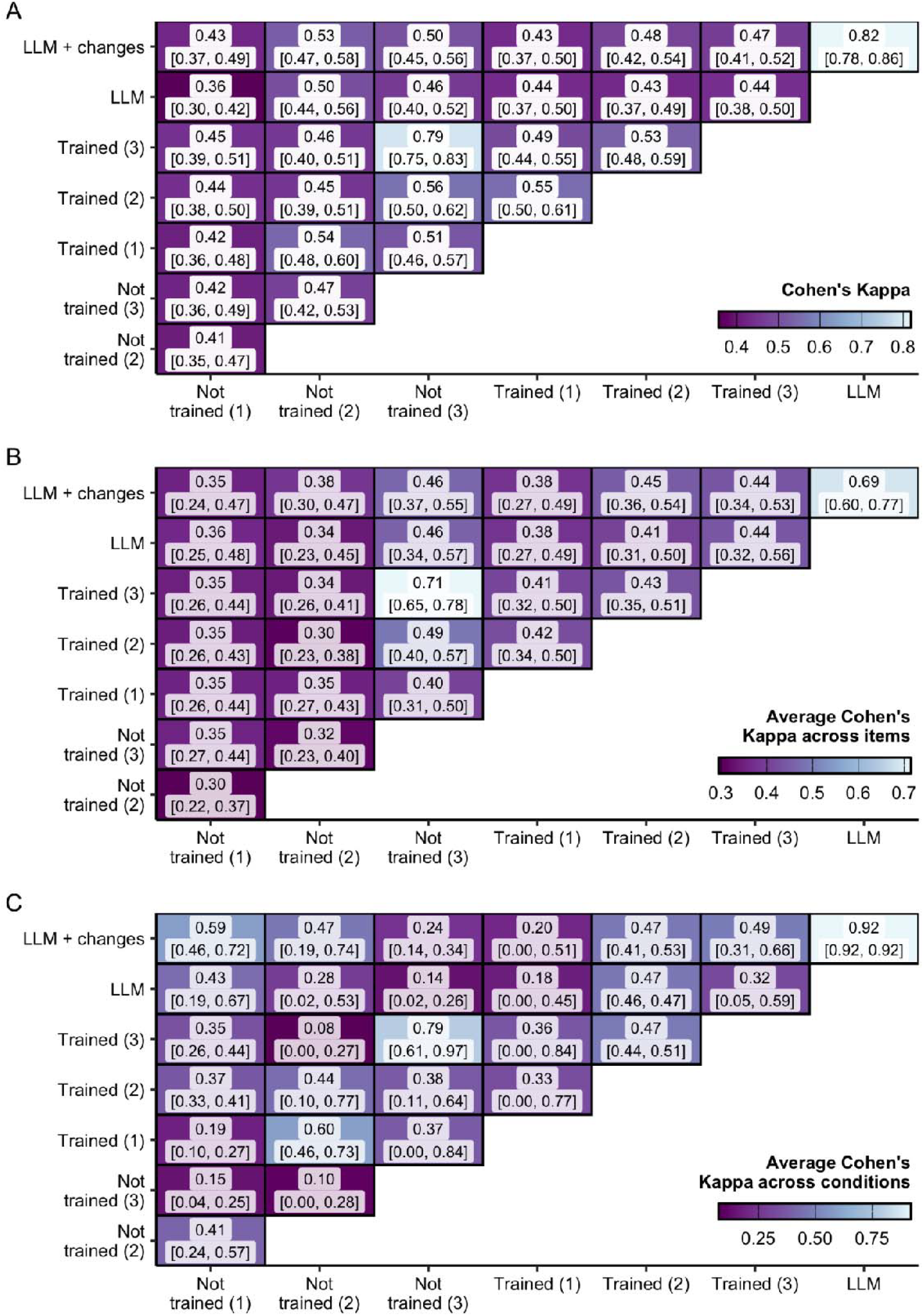
Inter-rater agreement as measured by Cohen’s Kappa between all human rater groups, the large language model (LLM) and the large language model upon iterative alterations seeking to improve the LLM performance Measured for Akinci D’Antonoli and others [17] A Cohen’s Kappa calculated using all items and Cohen’s Kappa/average Cohen’s Kappa. The text inside each cell represents Cohen’s Kappa/average Cohen’s Kappa and its 95% confidence interval in brackets.

When considering the scores between raters and the LLM, this picture is similar — routinely, the agreement between the LLM and raters is almost exclusively moderate. The single exception is that of the comparison between LLM and the rater group in the first experience level with no training in METRICS, where agreement is only fair (below 0.4). While lower in general, average Cohen’s Kappa calculated across all items between the LLM and rater groups are similar to those between other rater groups. Finally, we note that, for conditions, LLM-rater group comparisons tend to show lower inter-rater agreements (⊠=0.14 between the LLM and high expertise raters without training, CI 95%=[0.02, 0.26]). These are, however, not the lowest observed inter-rater agreements for METRICS conditions (⊠=0.08 between mid-expertise raters without training and high expertise raters with training, CI 95%=[0.00, 0.27], ⊠=0.10 between mid-expertise raters without training and high expertise raters without training, CI 95%=[0.00, 0.28]).

### Iteratively prompt enhancement

To understand and improve cases where LLM-human inter-rater agreement was low, we conducted a systematic assessment of the five items where the LLM showed the highest level of disagreement with human rater groups and observed the five scientific articles where this disagreement was most common. As illustrated in Figure 2, the 5 items with the highest disagreement are Item23, Item16, Item7, Item10 and Item19. Our structured output includes, for each field, a “reason” which highlights the logic behind the output of an LLM. Using this, we are able to understand each level of disagreement. In Supplementary Table 2 we present all of the analysed papers for each of the 5 mentioned items, and here we present short summaries of each:

**Figure 2.**
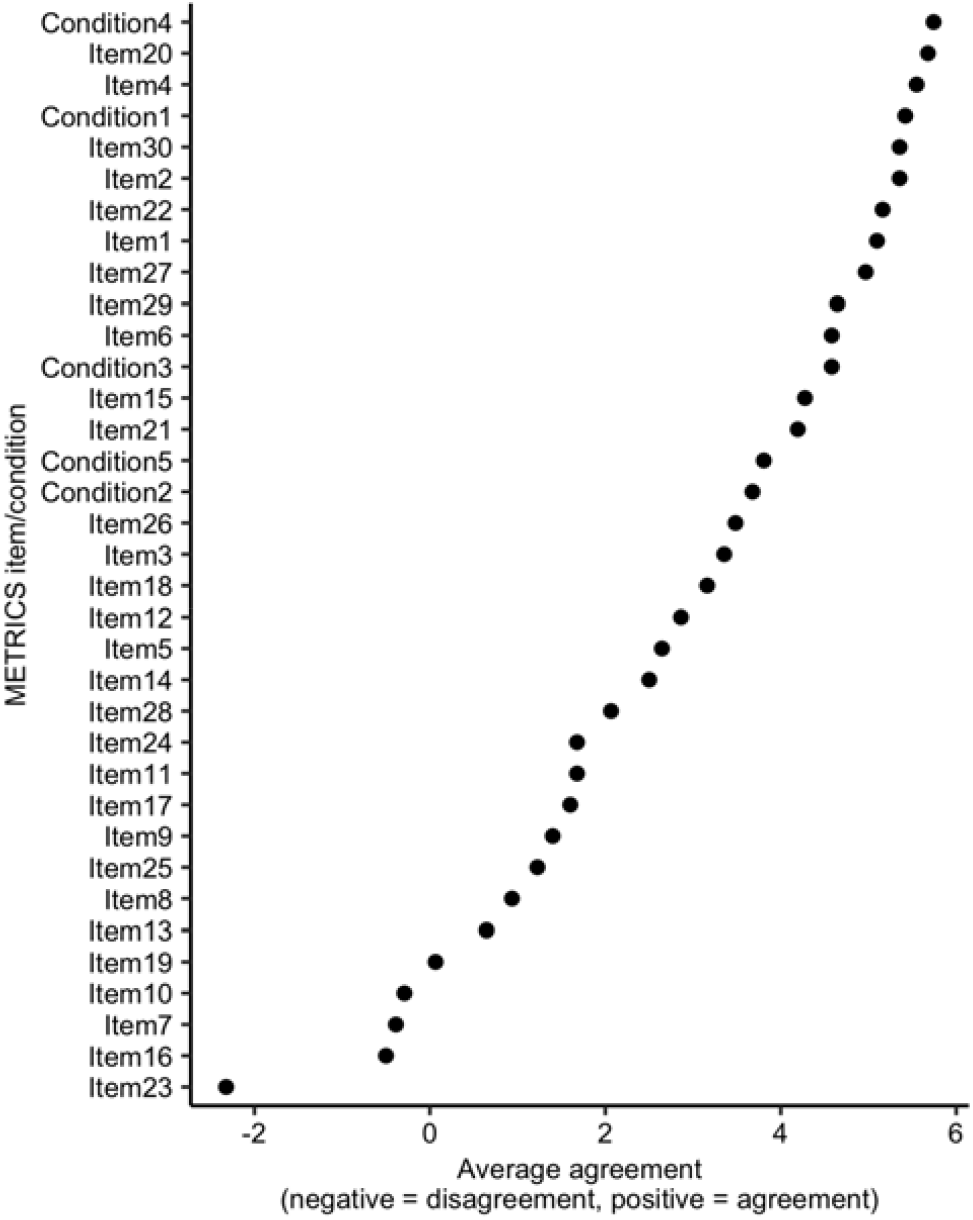
Average agreement between the LLM and all rater groups. Negative values imply a higher proportion of disagreements, whereas higher values imply a higher proportion of agreements.

- **Item 23 — Use of uni-parametric imaging or proof of its inferiority:** for this, the LLM mistakenly assumes that using multiple image transformations during radiomic feature extraction counts as “multi-parametric imaging”, or the LLM mistakenly interprets the paper and assumes that there are other categories apart from “uni-parametric imaging” and “multi-parametric imaging”. As an enhancement, we add to the prompt “Uni-parametric imaging implies the acquisition of a single imaging modality, as opposed to multi-parametric imaging, which involves the acquisition of multiple imaging modalities.”
- **Item 7: The interval between imaging used and reference standard:** we found in most instances, the interval between imaging and reference standard is acceptable (except for “Application of a comprehensive model based on CT radiomics and clinical features for postoperative recurrence risk prediction in non-small cell lung cancer”, where this interval is not mentioned) and follows the helping criteria in the METRICS webpage which we provide as part of the LLM prompt; as such, we attribute this to differences in research standards. As an enhancement, we add to the prompt “If there is no mention of when the reference standard was acquired relative to the diagnostic imaging exams, the answer should be”no”.”
- **Item 16 — Appropriateness of dimensionality compared to data size:** this point was frequently misinterpreted by the LLM. Indeed, the most common answer by the LLM was “yes”, while that of human rater groups was “no”. Given that there is no standard way of assessing this (i.e. there is no way of calculating whether data dimensionality is appropriate to dataset size by knowing about these two quantities alone), we cannot propose an enhancement to the prompt.
- **Item 10 — Test set segmentation masks produced by a single rater or automated tool:** here, the LLM considers a senior radiologist reviewing segmentations to be standard clinical practice, and given that the helper note provided in the prompt/METRICS tool website does not specifically address what “clinical practice” is, the LLM (very wrongfully) assumes that segmentation reviews are a standard part of it. As an enhancement, we add to the prompt “Keep in mind that having a radiologist review the results of the automated segmentation is not considered clinical practice.”
- **Item 19 — Handling of confounding factors:** while the helper note mentions that patient-specific confounders should be handled/discussed/removed, the LLM tends to ignore that these confounders should be patient-specific and sometimes focuses excessively on model- or acquisition-/scanner-specific confounders. **As an enhancement**, we add to the prompt “Be sure to focus on confounders stemming from the acquisition process, such as scanner type, manufacturer, or scanner parameters, or from the individual patient characteristics, such as age, weight, or comorbidities.”. To make the above point clearer, we note a few examples for this item where disagreements arose:
- In ‘A Radiomic “Warning Sign” of Progression on Brain MRI in Individuals with MS’, the authors offer an analysis of a pipeline-specific confounder which could affect how predictions were obtained or are interpreted. However, this confounder does not represent anything inherent to the patient or to the acquisition process, but rather to the algorithm developed during this work. All raters answered “no” while the LLM answered “yes”.
- In “CNN-based multi-modal radiomics analysis of pseudo-CT utilization in MRI-only brain stereotactic radiotherapy: a feasibility study”, the authors do offer an analysis of how MRI contrast presence affects model performance. All raters answered “no” while the LLM answered “yes”.
- In the remaining articles where the LLM answered “yes” while rater groups tended towards “no”, there are some analysed factors which can be considered confounders but it is understandable that they are not necessarily regarded as relevant in modern clinical practice.

With these prompt enhancements, we achieve a consistent improvement across the overall, item average and condition average Cohen’s Kappa (Figure 1). However, we note that these improvements, albeit recurrent, are not statistically significant (i.e. there is significant interval overlap). Calculating the average Cohen’s Kappa for the five items targeted by our enhancements shows, however, that the enacted changes led to improved inter-rater agreement between the LLM and the human rater groups for 4 out of 6 inter-rater agreements (Figure 3).

**Figure 3.**
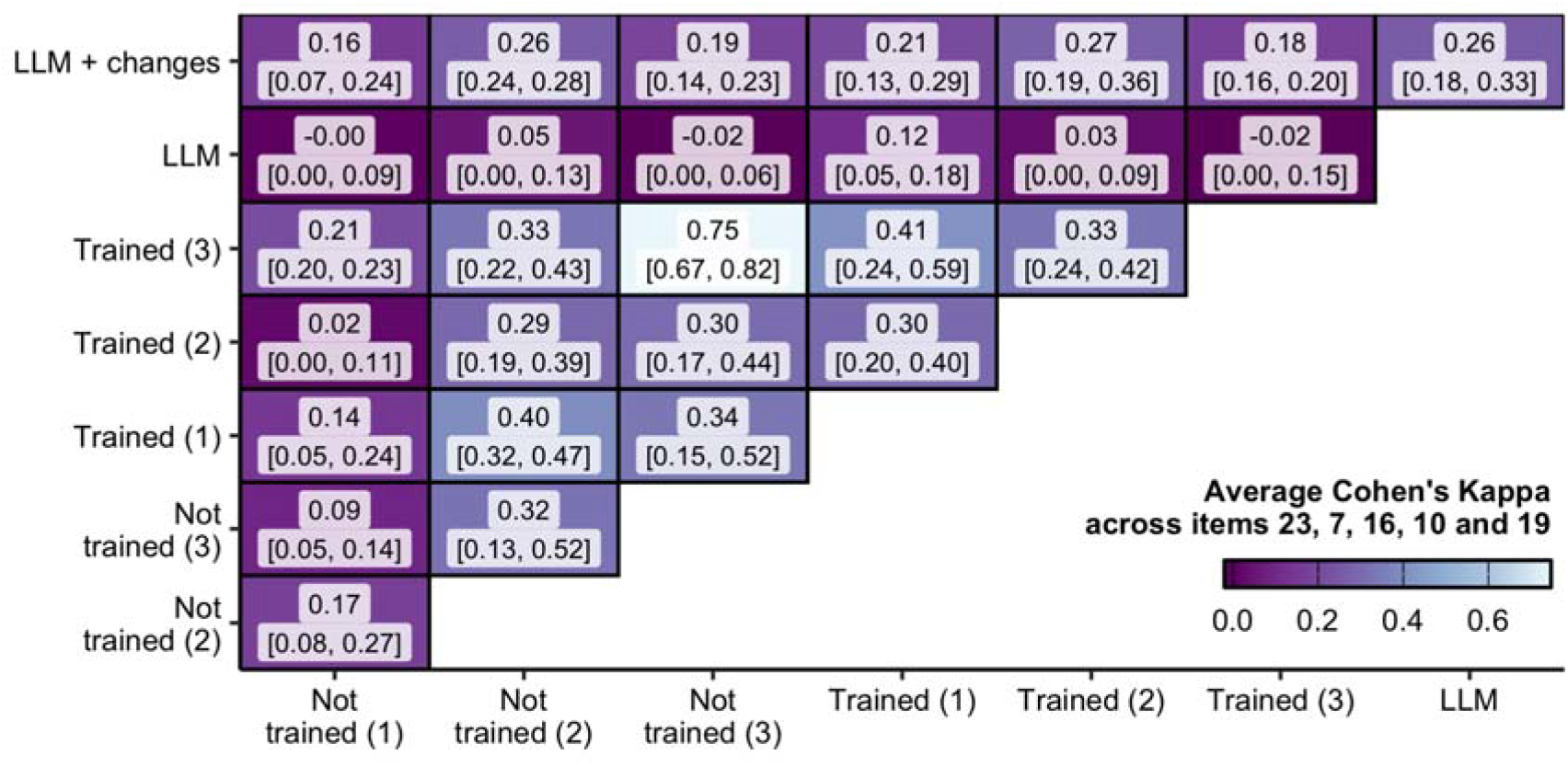
Inter-rater agreement as measured by the average Cohen’s Kappa for items 23, 7, 16, 10 and 19 between all human rater groups, the large language model (LLM) and the large language model upon iterative alterations seeking to improve the LLM performance. Measured for Akinci D’Antonoli and others (Akinci D’Antonoli et al. 2025).

Finally, we calculated for all human and LLM raters the METRICS score. We then assessed the relative and absolute differences between all raters. In Figure 4 and Figure 5, we show that these differences are small or non-existent and comparable between one another. As noted earlier, these differences highlight greater similarities between high-expertise raters with and without training while remaining relatively between one another. To better formalise these comparisons, we perform an analysis of variance (ANOVA) where the METRICS score is the dependent variable and a categorical annotation denoting each human/LLM rater group. An F-test denoted a significant difference between the category averages (p<0.001), and further post-hoc Tukey Honest Significant Difference tests showed that 40% of comparisons between human rater groups were statistically significant (6/15), whereas 58% comparisons between human reader groups and LLMs were statistically significant (7/12), which constitutes a statistically insignificant difference between proportions (p=0.56 for a two-sample two-way test for equality of proportions).

**Figure 4.**
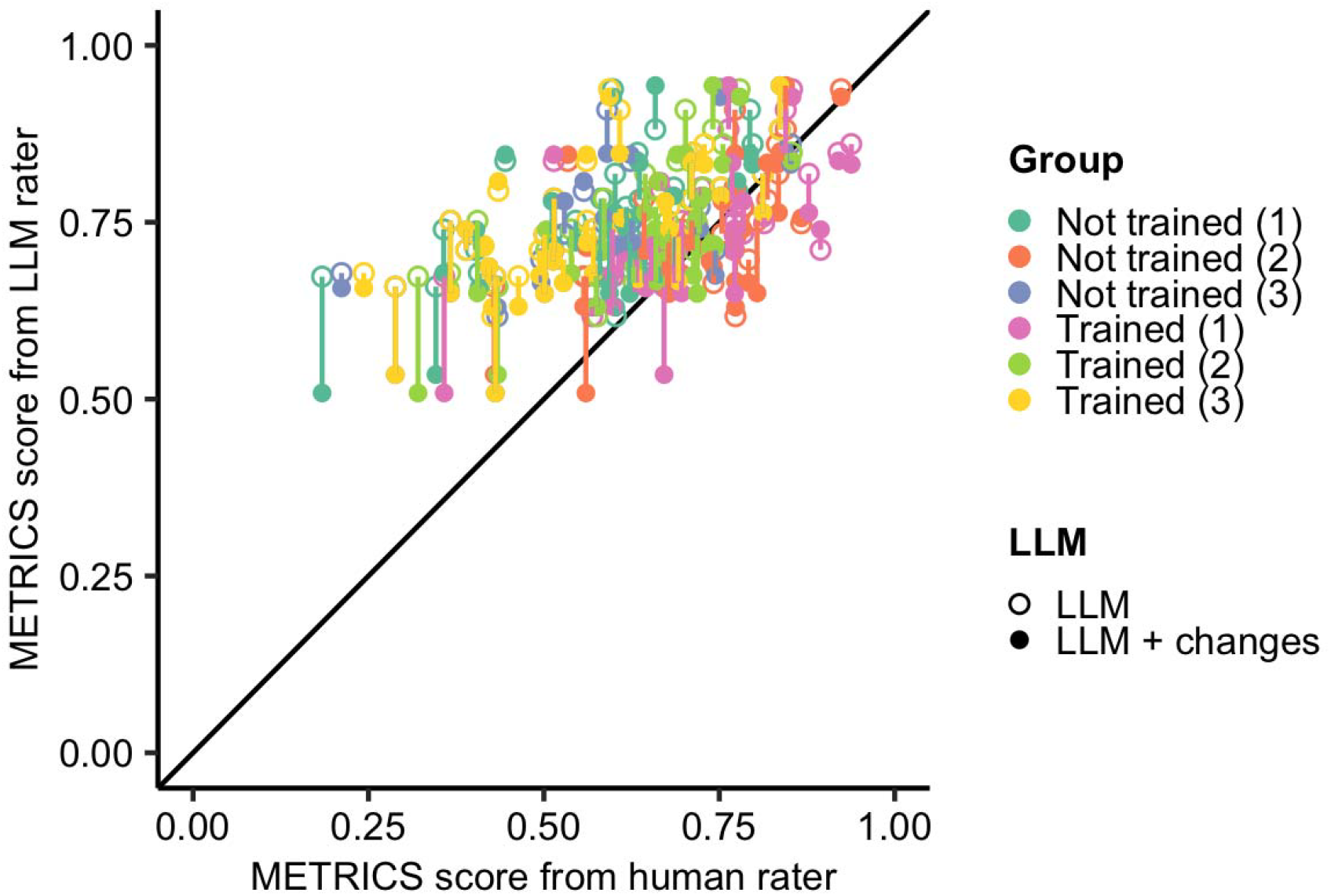
Comparison of METRICS score between all human rater groups, the large language model (LLM) and LLM outputs for Akinci D’Antonoli and others [17]. Colours represent different human rater groups, and shapes represent whether the used LLM was with or without changes (empty circle and full circle, respectively). The black diagonal line represents the identity.

**Figure 5.**
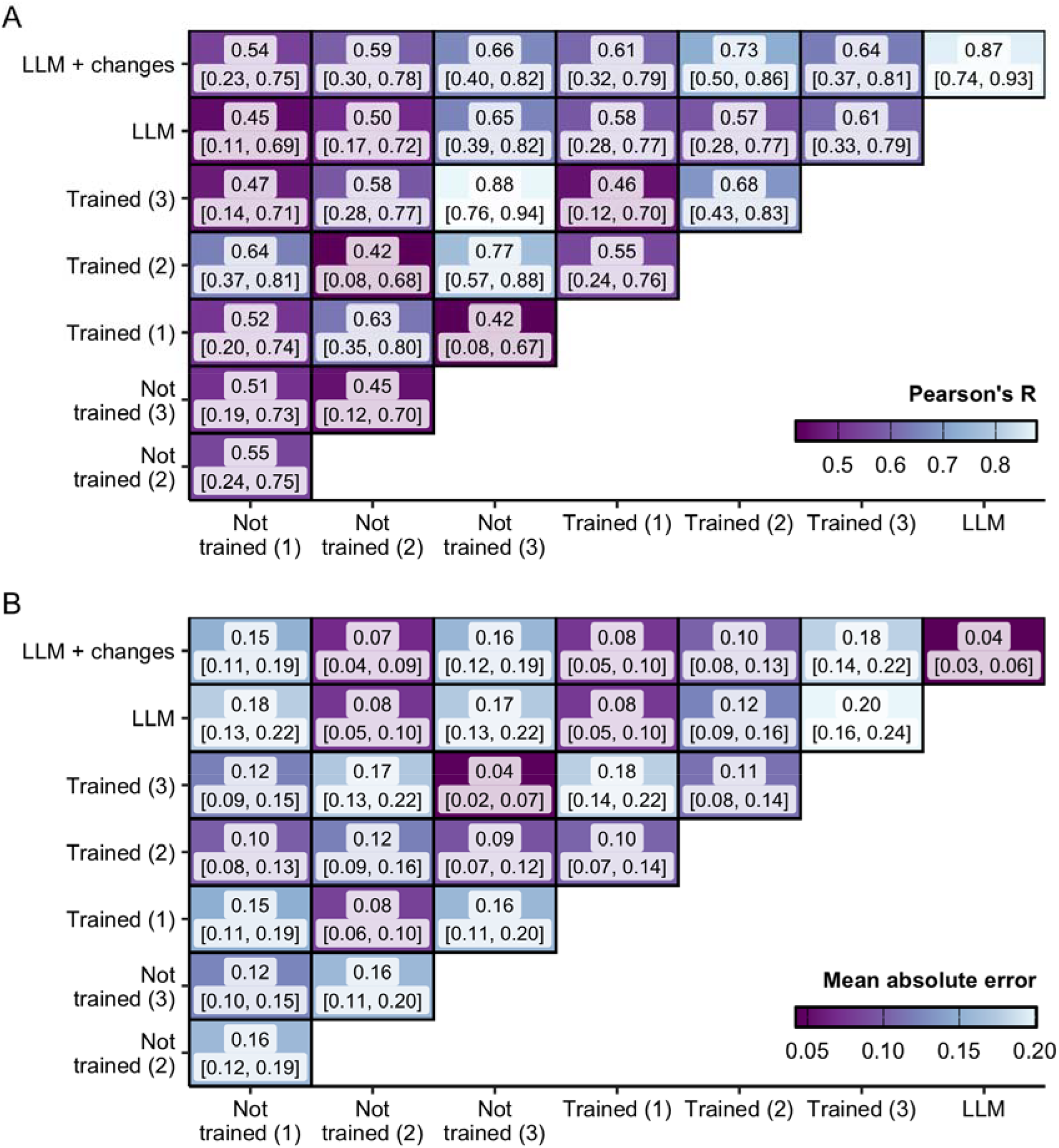
METRICS score Pearons’s R (A) and mean absolute error (B) between all human rater groups, the large language model (LLM) and the large language model upon iterative alterations seeking to improve the LLM performance. Measured for Akinci D’Antonoli and others [17]. The text inside each cell represents each metric and its 95% confidence interval in brackets.

**Figure 6.**
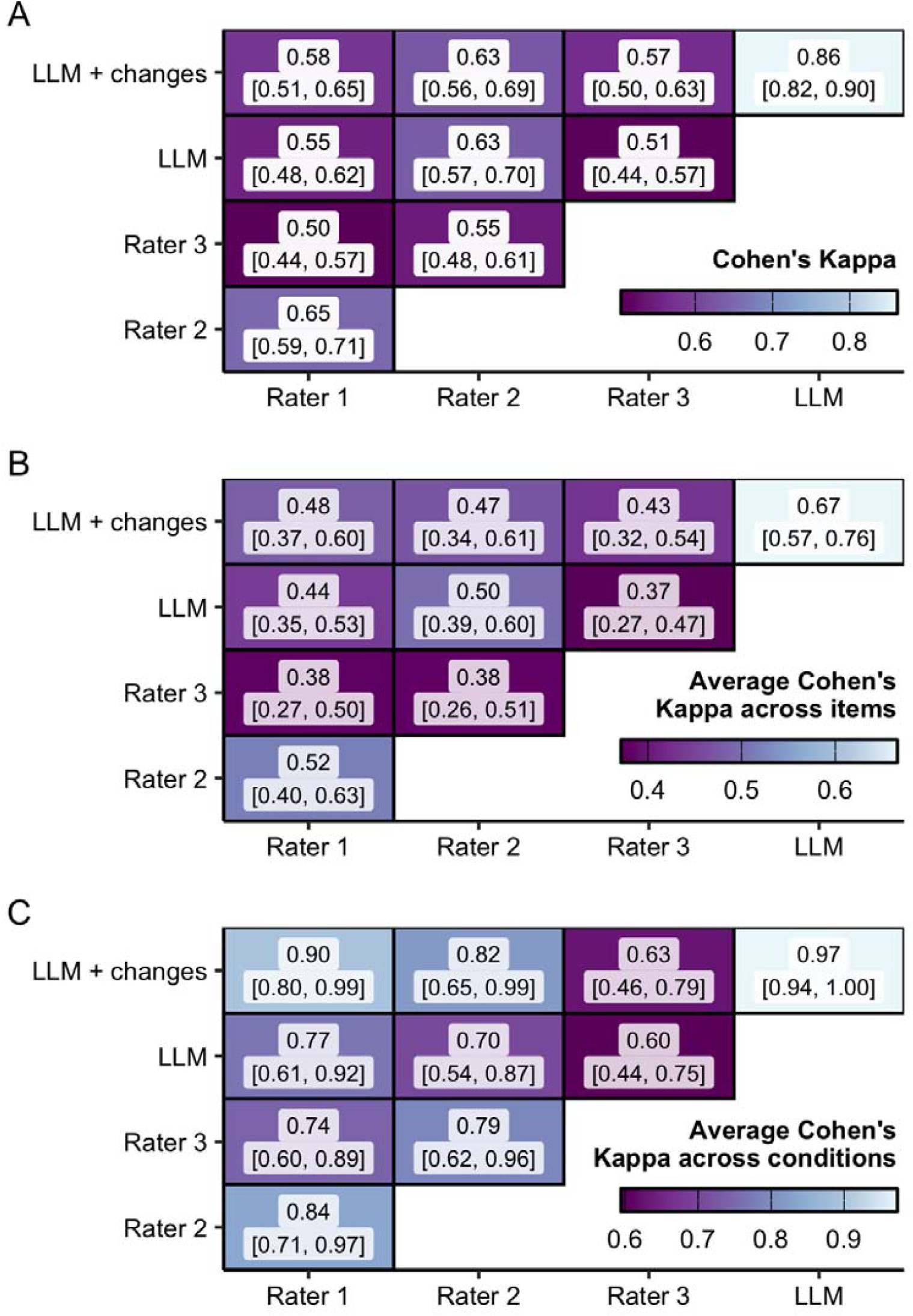
Inter-rater agreement as measured by Cohen’s Kappa between all human rater groups, the large language model (LLM) and the large language model upon iterative alterations seeking to improve the LLM performance. Measured for Kocak and others [16]. A. Cohen’s Kappa calculated using all items and conditions. B. Average item Cohen’s Kappa. C. Average condition Cohen’s Kappa. For A-C, colours represent the Cohen’s Kappa/average Cohen’s Kappa. The text inside each cell represents Cohen’s Kappa/average Cohen’s Kappa and its 95% confidence interval in brackets.

### Generalisation of conclusions to other surveys

To better understand whether these results were not chance findings and to further validate whether our prompt enhancement protocol is useful within datasets, we reproduced our analysis on Kocak and others (2025) [16]. We show good inter-rater agreement between the LLM with and without changes and all raters. However, we note that — while there is a positive trend — no statistically significant changes were found between the LLM and the LLM + changes. Similarly to what was performed for Akinci D’Antonoli, we calculated the METRICS scores for all human and LLM raters. Following a statistically significant F-score for a linear model associating the METRICS score with a category for each rater (p<0.001), 33% (1/3) and 66% (4/6) of post-hoc tests were statistically significant for human-human comparisons and for human-LLM comparisons, respectively (p=0.8 for a two-sample two-way test for equality of proportions).

## Discussion

Here, we show that an automatic assessment of METRICS using a commercially available LLM is a feasible complement to those provided by radiologists when assessing the scientific quality of radiomics publications.

The topic of measuring the quality of scientific production is not without controversy. Most of the well-known measures focus on the productivity of scientists (i.e. impact factors or h-indices [25]) rather than on the quality of individual publications. Similar metrics can be used retrospectively for scientific articles — indeed, one can ascribe importance to a scientific article which has accrued a large number of citations or, in other words, a high impact factor. However, this is not a measure of the scientific quality of the paper itself. Importantly, it is not a measure of the methodological quality of the article. Some fields — such as evidence-based medicine — focus on categorizing the different levels of evidence which can be raised to support a claim [26]. For instance, a randomised control trial provides stronger evidence for an effect than a case-control study.

However, most radiomics studies are retrospective cohort/case-control studies, placing them roughly at the same evidence level. Other tools focus on analysing systematic or meta-reviews [27], but the point at which these reviews can be considered may have already signified a significant amount of investment into a specific and overly optimistic solution. When performing meta-studies more broadly, individual publications can be assessed using a number of different guidelines [28]. Indeed, this field is abundant with different metrics and METRICS itself does appear to take inspiration from some of them: concerns for sample size, potential confounders, accurate description of methodology and sufficient clarification that allows for the replication of methods [29,30]. METRICS thus serves a unique and relevant purpose by combining and introducing radiomics-specific items to these sorts of assessments. Together with RQS [13] or other radiomics assessments focusing on radiomic feature repeatability/reproducibility [31], it stands as one of the few instruments for researchers wishing to assess the quality of their radiomics research.

However, it should be noted that there appear to be systematic pitfalls: the fair-to-moderate inter-rater agreement, as well as the notorious complexity arising from specific items. Item 16, concerning the adequacy of the sample size when compared with the dimensionality of the data, was raised as a METRICS item which was hard to understand by radiologists [17]. Our analysis here showed that this issue extends to LLM-based assessments. Nonetheless, it should be noted that RQS leads to even lower inter-rater reliability [32].

The literature using and assessing LLMs for scientific activities has been on the rise. Here, we particularly note that this is also the case for LLMs for scientific evaluation. For the more open-ended task of reviewing scientific articles, approaches can be clustered into a few trends. Some works have shown that the standard use of LLMs for peer-reviewing scientific articles leads to good results [21,33]. Others have developed tool-based approaches which extract and mine papers similar to a given article to confirm that the research is consistent with previous works [34–37]. An additional work even used an iterative LLM-based approach to produce a questionnaire which reviewers have to follow to review a given paper, using that questionnaire to prompt a separate LLM for a review and improving the usefulness of the review [38]. Importantly, a recent preprint has shown that LLMs can be used — by simply providing feedback on reviews — make reviews more informative [23]. Auto-METRICS here adds to this body of research, showing that standardized automated assessments of methodological quality are not impossible in radiomics research.

We note this work does not come without its **Limitations**. While this is a demonstration of the potential of LLMs for scientific methodology assessment, we note that the dataset size is relatively small, requiring larger sample sizes for better assessment. Additionally, different LLM-based strategies or prompts might lead to different results — famously, chain of thought prompting, where an LLM is prompted to think carefully before producing an output, leads to better results [39]. However, we note that Auto-METRICS results are already relatively good as far as inter-rater agreements are concerned. Finally, this work should not be seen as an incentive for indiscriminate use of LLMs in scientific writing or peer-review. Indeed, in scientific writing in general, LLMs have been increasingly used [40], with LLM use often going undisclosed in many scenarios [41]. Furthermore, reviewers who are less confident in their evaluations and submitting their reviews closer to deadlines were found to be more likely to contribute with text associated with LLM editing in artificial intelligence conferences [42].

## Data Availability

All data produced are available online at https://github.com/josegcpa/auto-metrics.

## Supplementary information

### Supplementary methods

#### Prompt used for Auto-METRICS

\# *Instructions*

*You are an expert radiologist with decades of experience in developing and implementing clinical artificial intelligence*.

*You have to offer ratings to a scientific article of clinical importance at the end of this prompt according to a set of criteria*.

*You must answer carefully and thoughtfully, considering the context of the article and your expertise*.

*Be extremely thorough and conservative with your answers as these tools are supposed to be deployed in the clinic*.

\# *Evaluation*

*## Metrics definition*

*There are 3*0 *criteria in total and each criterion is grouped into one of 9 categories*.

*There are, additionally, 5 conditions which define whether some criterion should be filled or not*.

*The 5 Conditions are defined below under “## Conditions”. A short explanation is provided for each*.

*The 30 criteria are defined below under “## Criteria”. Each is grouped under its respective category. A short explanation is provided for each*.

\## *Rating Rubric*

*No: no evidence that this criterion is being followed in this publication*

*Yes: evidence that this criterion is being followed in this publication*

*n/a: not applicable*

*Reason: a short explanation for each ranking*

*# Input format*

*The article text is provided below under “# Article text”. Anything outside of this text should not be evaluated*.

*# Output format*

*You have to output:*

- *a summary of the article accurately representing the main conclusion of the article*
- *the answers for conditions (Yes, No) and a short explanation for your decision*
- *the ratings (Yes, No or n/a) for each criterion and a short explanation for your decision*

### Supplementary tables

**Supplementary Table 1:**
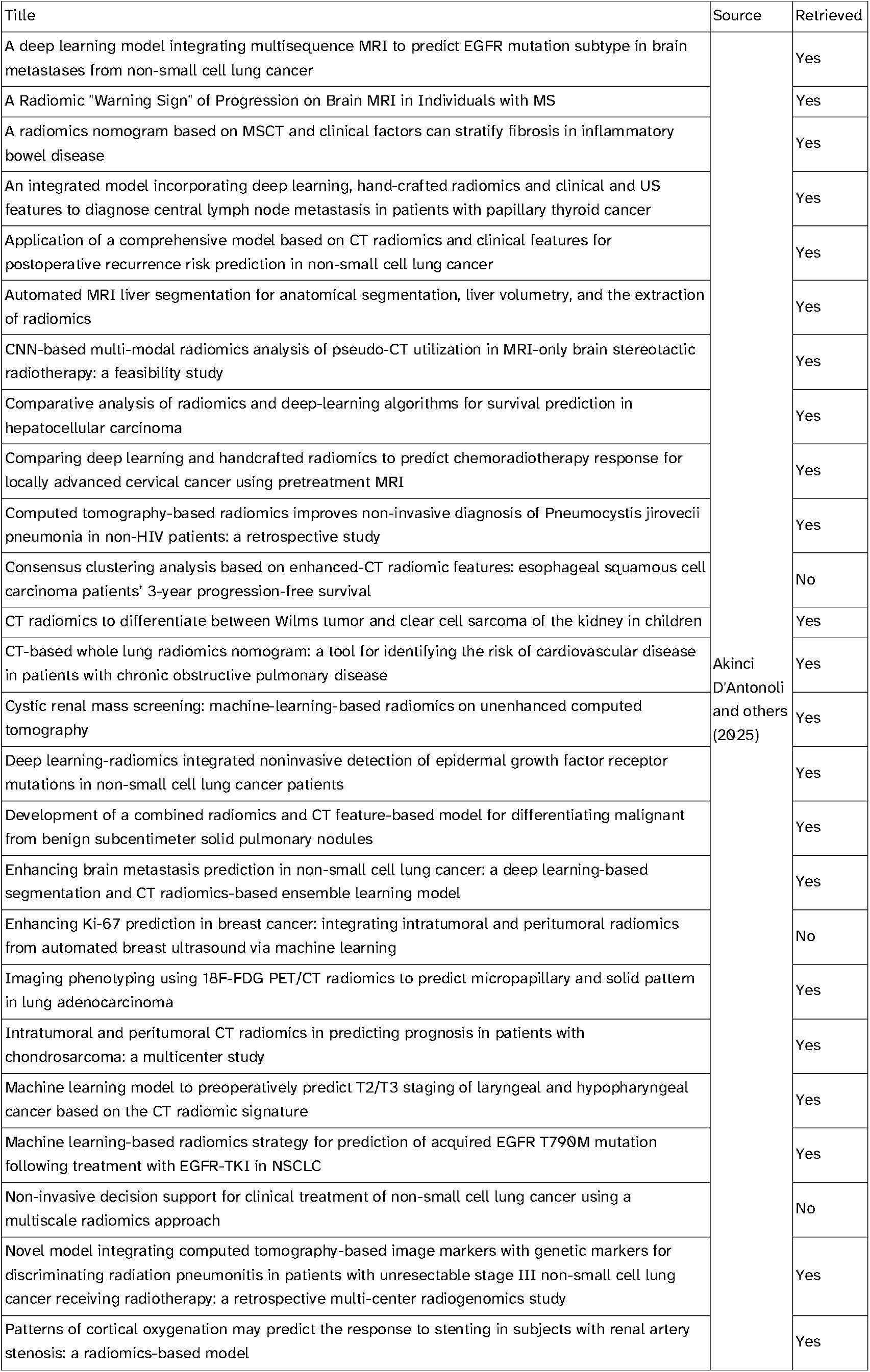
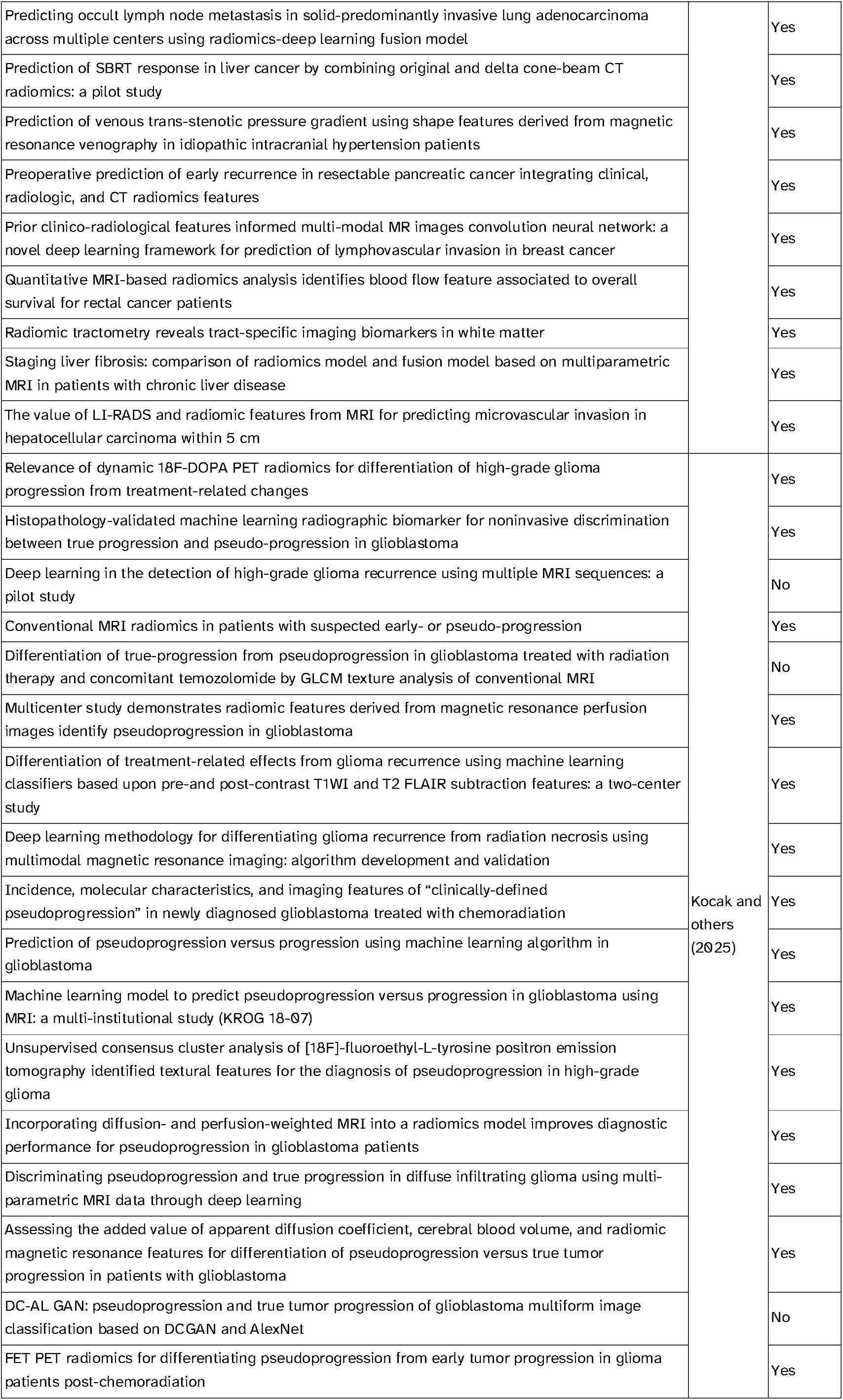
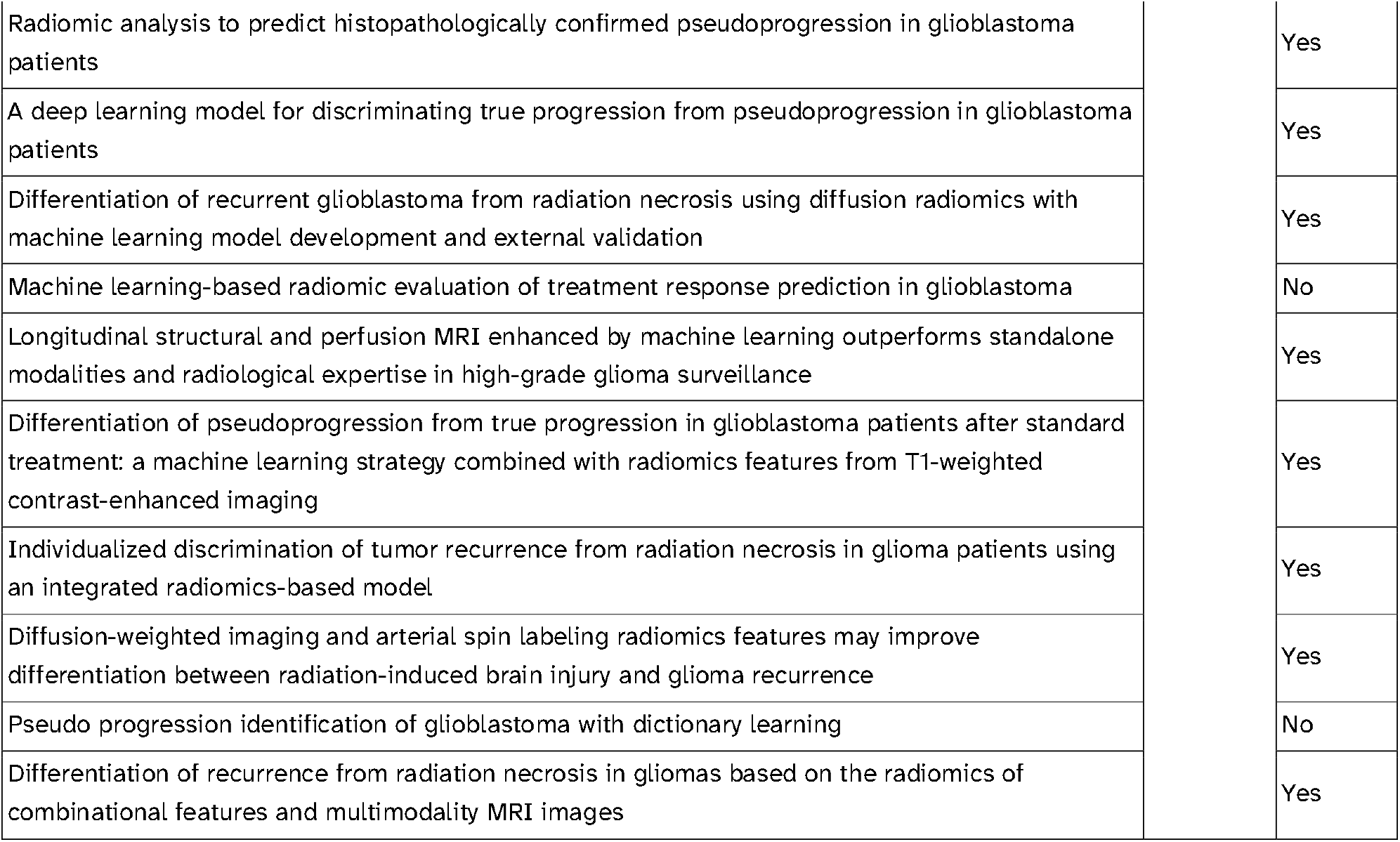
list of articles analysed in [17] and [16] and whether they were retrieved or not.

**Supplementary Table 2:**
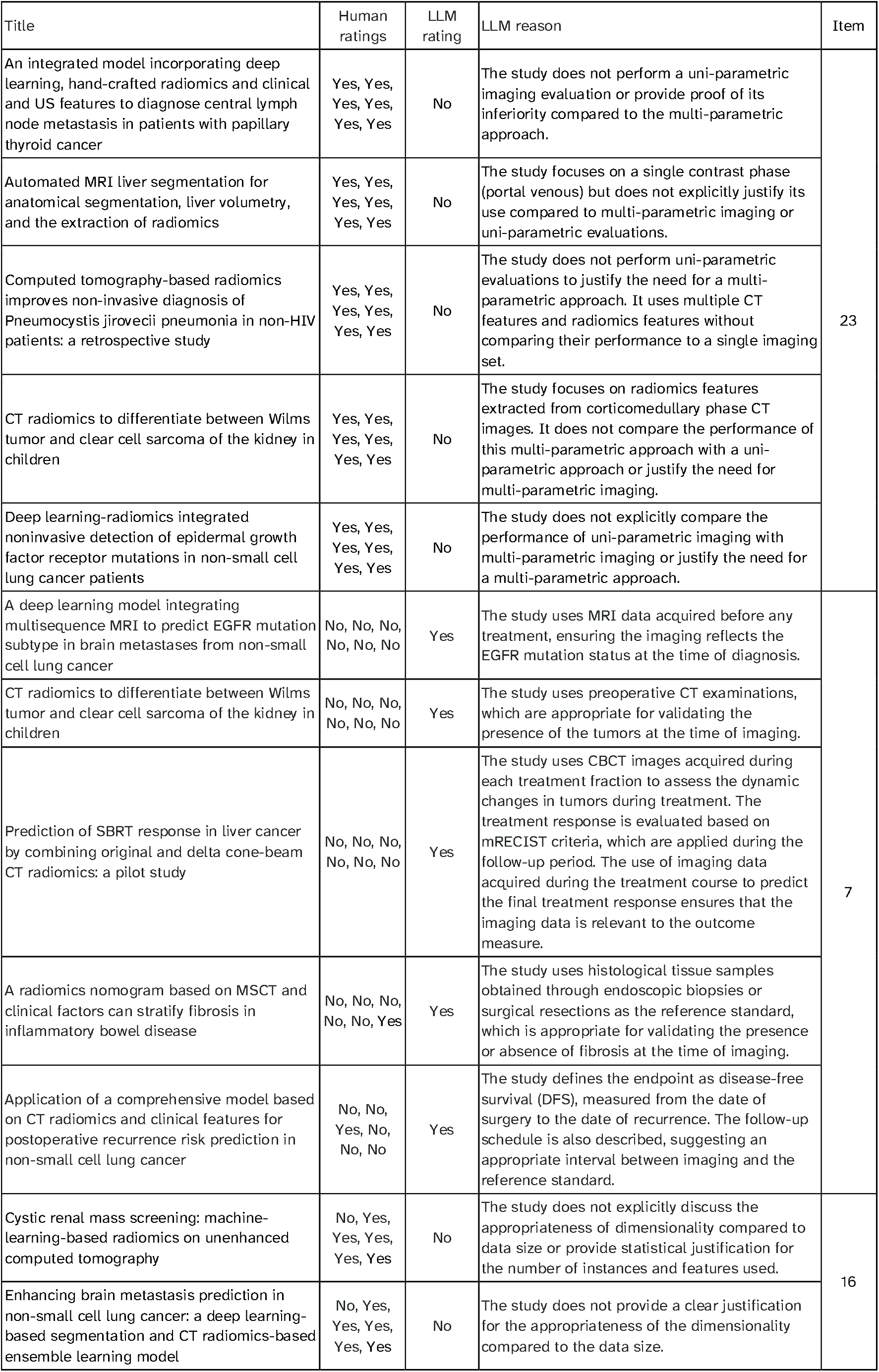
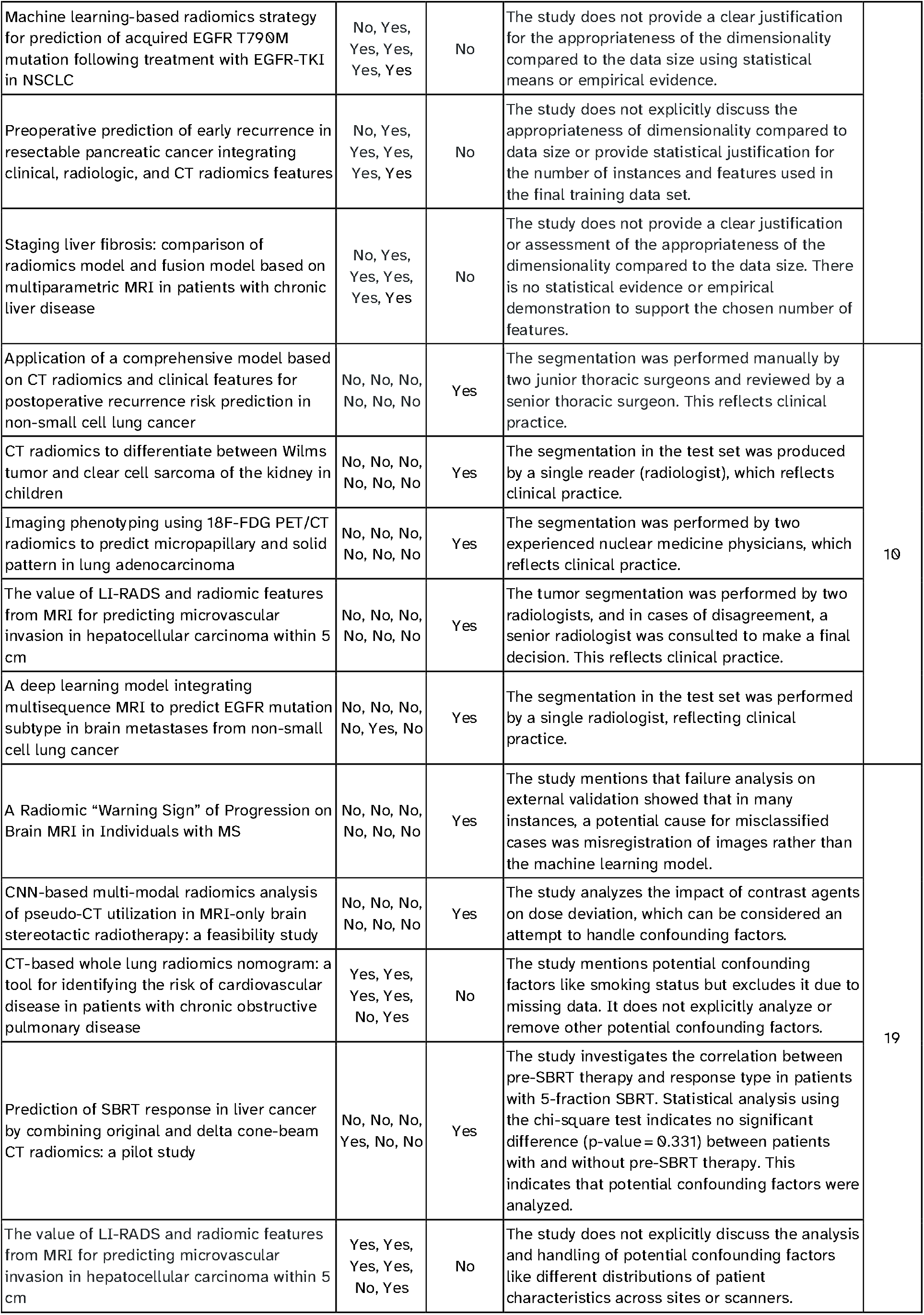
List of large language model (LLM) errors for 5 papers with high disagreement levels across items with high average disagreements.

## References

1. Kapoor S, Narayanan A. Leakage and the reproducibility crisis in machine-learning-based science. Patterns (N Y). 2023;4:100804.

2. AAAI 2025 Presidential Panel on the Future of AI Research. In: AAAI [Internet]. 28 Feb 2025 [cited 7 Apr 2025]. Available: https://aaai.org/about-aaai/presidential-panel-on-the-future-of-ai-research/

3. Maleki F, Ovens K, Gupta R, Reinhold C, Spatz A, Forghani R. Generalizability of Machine Learning models: Quantitative evaluation of three methodological pitfalls. Radiol Artif Intell. 2023;5:e220028.

4. Kocak B, Bulut E, Bayrak ON, Okumus AA, Altun O, Borekci Arvas Z, et al. NEgatiVE results in Radiomics research (NEVER): A meta-research study of publication bias in leading radiology journals. Eur J Radiol. 2023;163:110830.

5. Lu L, Ahmed FS, Akin O, Luk L, Guo X, Yang H, et al. Uncontrolled confounders may lead to false or overvalued radiomics signature: A proof of concept using survival analysis in a multicenter cohort of kidney cancer. Front Oncol. 2021;11:638185.

6. Cannella R, Santinha J, Bèaufrere A, Ronot M, Sartoris R, Cauchy F, et al. Performances and variability of CT radiomics for the prediction of microvascular invasion and survival in patients with HCC: a matter of chance or standardisation? Eur Radiol. 2023;33:7618– 7628.

7. Demircioğlu A. Measuring the bias of incorrect application of feature selection when using cross-validation in radiomics. Insights Imaging. 2021;12:172.

8. Berenguer R, Pastor-Juan M del R, Canales-Vázquez J, Castro-García M, Villas MV, Mansilla Legorburo F, et al. Radiomics of CT features may be nonreproducible and redundant: Influence of CT acquisition parameters. Radiology. 2018;288:407–415.

9. Gillies RJ, Kinahan PE, Hricak H. Radiomics: Images are more than pictures, they are data. Radiology. 2016;278:563–577.

10. Pinto Dos Santos D, Dietzel M, Baessler B. A decade of radiomics research: are images really data or just patterns in the noise? Eur Radiol. 2021;31:1–4.

11. Horvat N, Papanikolaou N, Koh D-M. Radiomics beyond the hype: A critical evaluation toward oncologic clinical use. Radiol Artif Intell. 2024;6:e230437.

12. Moskowitz CS, Welch ML, Jacobs MA, Kurland BF, Simpson AL. Radiomic analysis: Study design, statistical analysis, and other bias mitigation strategies. Radiology. 2022;304:265–273.

13. Park JE, Kim HS, Kim D, Park SY, Kim JY, Cho SJ, et al. A systematic review reporting quality of radiomics research in neuro-oncology: toward clinical utility and quality improvement using high-dimensional imaging features. BMC Cancer. 2020;20:29.

14. Kocak B, Akinci D’Antonoli T, Mercaldo N, Alberich-Bayarri A, Baessler B, Ambrosini I, et al. METhodological RadiomICs Score (METRICS): a quality scoring tool for radiomics research endorsed by EuSoMII. Insights Imaging. 2024;15:8.

15. Koçak B, Akinci D’Antonoli T, Cuocolo R. Exploring radiomics research quality scoring tools: a comparative analysis of METRICS and RQS. Diagn Interv Radiol. 2024;30:366– 369.

16. Kocak B, Mese I, Ates Kus E. Radiomics for differentiating radiation-induced brain injury from recurrence in gliomas: systematic review, meta-analysis, and methodological quality evaluation using METRICS and RQS. Eur Radiol. 2025; 1–16.

17. Akinci D’Antonoli T, Cavallo AU, Kocak B, Borgheresi A, Ponsiglione A, Stanzione A, et al. Reproducibility of methodological radiomics score (METRICS): an intra- and inter-rater reliability study endorsed by EuSoMII. Eur Radiol. 2025; 1–13.

18. Zheng Y, Koh HY, Ju J, Nguyen ATN, May LT, Webb GI, et al. Large language models for scientific discovery in molecular property prediction. Nat Mach Intell. 2025;7:437–447.

19. Luo X, Rechardt A, Sun G, Nejad KK, Yáñez F, Yilmaz B, et al. Large language models surpass human experts in predicting neuroscience results. Nat Hum Behav. 2025;9:305–315.

20. Evans J, D’Souza J, Auer S. Large Language Models as evaluators for scientific synthesis. arXiv [cs.CL]. 2024. Available: http://arxiv.org/abs/2407.02977

21. Liang W, Zhang Y, Cao H, Wang B, Ding DY, Yang X, et al. Can large language models provide useful feedback on research papers? A large-scale empirical analysis. NEJM AI. 2024;1. doi:10.1056/aioa2400196

22. Cao C, Sang J, Arora R, Kloosterman R, Cecere M, Gorla J, et al. Prompting is all you need: LLMs for systematic review screening. Health Informatics. medRxiv; 2024. Available: https://www.medrxiv.org/content/10.1101/2024.06.01.24308323v1

23. Thakkar N, Yuksekgonul M, Silberg J, Garg A, Peng N, Sha F, et al. Can LLM feedback enhance review quality? A randomized study of 20K reviews at ICLR 2025. arXiv [cs.AI]. 2025. Available: http://arxiv.org/abs/2504.09737

24. Generate structured output with the Gemini API. In: Google AI for Developers [Internet]. [cited 7 Apr 2025]. Available: https://ai.google.dev/gemini-api/docs/structured-output?lang=python

25. Kumar M. Evaluating scientists: Citations, impact factor, h-index, online page hits and what else? IETE Tech Rev. 2009;26:165.

26. Koretz RL. Assessing the evidence in evidence-based medicine. Nutr Clin Pract. 2019;34:60–72.

27. Lunny C, Kanji S, Thabet P, Haidich A-B, Bougioukas KI, Pieper D. Assessing the methodological quality and risk of bias of systematic reviews: primer for authors of overviews of systematic reviews. BMJ Med. 2024;3:e000604.

28. Luchini C, Veronese N, Nottegar A, Shin JI, Gentile G, Granziol U, et al. Assessing the quality of studies in meta-research: Review/guidelines on the most important quality assessment tools. Pharm Stat. 2021;20:185–195.

29. Vandenbroucke JP, von Elm E, Altman DG, Gøtzsche PC, Mulrow CD, Pocock SJ, et al. Strengthening the Reporting of Observational Studies in Epidemiology (STROBE): explanation and elaboration. PLoS Med. 2007;4:e297.

30. Liu X, Cruz Rivera S, Moher D, Calvert MJ, Denniston AK, SPIRIT-AI and CONSORT-AI Working Group. Reporting guidelines for clinical trial reports for interventions involving artificial intelligence: the CONSORT-AI extension. Nat Med. 2020;26:1364–1374.

31. Pfaehler E, Zhovannik I, Wei L, Boellaard R, Dekker A, Monshouwer R, et al. A systematic review and quality of reporting checklist for repeatability and reproducibility of radiomic features. Phys Imaging Radiat Oncol. 2021;20:69–75.

32. Akinci D’Antonoli T, Cavallo AU, Vernuccio F, Stanzione A, Klontzas ME, Cannella R, et al. Reproducibility of radiomics quality score: an intra- and inter-rater reliability study. Eur Radiol. 2024;34:2791–2804.

33. Robertson Z. GPT4 is slightly helpful for peer-review assistance: A pilot study. arXiv [cs.HC]. 2023. Available: http://arxiv.org/abs/2307.05492

34. Wang Q, Zeng Q, Huang L, Knight K, Ji H, Rajani NF. ReviewRobot: Explainable paper review generation based on knowledge synthesis. arXiv [cs.CL]. 2020. Available: http://arxiv.org/abs/2010.06119

35. Chamoun E, Schlichktrull M, Vlachos A. Automated focused feedback generation for scientific writing assistance. arXiv [cs.CL]. 2024. Available: http://arxiv.org/abs/2405.20477

36. D’Arcy M, Hope T, Birnbaum L, Downey D. MARG: Multi-Agent Review Generation for Scientific Papers. arXiv [cs.CL]. 2024. Available: http://arxiv.org/abs/2401.04259

37. Yu J, Ding Z, Tan J, Luo K, Weng Z, Gong C, et al. Automated peer reviewing in paper SEA: Standardization, evaluation, and Analysis. arXiv [cs.CL]. 2024. Available: http://arxiv.org/abs/2407.12857

38. Gao Z, Brantley K, Joachims T. Reviewer2: Optimizing Review Generation Through Prompt Generation. arXiv [cs.CL]. 2024. Available: http://arxiv.org/abs/2402.10886

39. Wei J, Wang X, Schuurmans D, Bosma M, Ichter B, Xia F, et al. Chain-of-thought prompting elicits reasoning in large language models. arXiv [cs.CL]. 2022. Available: http://arxiv.org/abs/2201.11903

40. Liang W, Zhang Y, Wu Z, Lepp H, Ji W, Zhao X, et al. Mapping the increasing use of LLMs in scientific papers. arXiv [cs.CL]. 2024. Available: http://arxiv.org/abs/2404.01268

41. Lazebnik T, Rosenfeld A. Detecting LLM-assisted writing in scientific communication: Are we there yet? J Data Inf Sci. 2024;9:4–13.

42. Liang W, Izzo Z, Zhang Y, Lepp H, Cao H, Zhao X, et al. Monitoring AI-modified content at scale: A case study on the impact of ChatGPT on AI conference peer reviews. arXiv [cs.CL]. 2024. Available: http://arxiv.org/abs/2403.07183

